# Time of day of vaccination affects SARS-CoV-2 antibody responses in an observational study of healthcare workers

**DOI:** 10.1101/2021.10.28.21265499

**Authors:** Wei Wang, Peter Balfe, David W Eyre, Sheila F Lumley, Denise O’Donnell, Fiona Warren, Derrick W Crook, Katie Jeffery, Philippa C Matthews, Elizabeth B Klerman, Jane A McKeating

**Affiliations:** Division of Sleep and Circadian Disorders, Brigham and Women’s Hospital; Division of Sleep Medicine, Harvard Medical School, US; Nuffield Department of Medicine, University of Oxford, Oxford, UK; Big Data Institute, Nuffield Department of Population Health, University of Oxford, Oxford, UK; Oxford University Hospitals NHS Foundation Trust, John Radcliffe Hospital, Oxford, UK; NIHR Oxford Biomedical Research Centre, University of Oxford, Oxford, OX3 9DU, UK; Radcliffe Department of Medicine, University of Oxford, UK; Department of Neurology, Massachusetts General Hospital; Division of Sleep and Circadian Disorders, Brigham and Women’s Hospital; Division of Sleep Medicine, Harvard Medical School, US; Chinese Academy of Medical Sciences Oxford Institute, University of Oxford, Oxford, UK

## Abstract

The COVID-19 pandemic caused by severe acute respiratory syndrome coronavirus 2 (SARS-CoV-2) is a global crisis with unprecedented challenges for public health. Vaccinations against SARS-CoV-2 have slowed the incidence of new infections and reduced disease severity. As the time-of-day of vaccination has been reported to influence host immune responses to multiple pathogens, we quantified the influence of SARS-CoV-2 vaccination time, vaccine type, age, sex, and days post-vaccination on anti-Spike antibody responses in healthcare workers. The magnitude of the anti-Spike antibody response associated with the time-of-day of vaccination, vaccine type, participant age, sex, and days post vaccination. These results may be relevant for optimizing SARS-CoV-2 vaccine efficacy.

---

The circadian clock is an endogenous 24 hour clock that regulates many aspects of physiology, including the response to infectious disease and vaccination (Allada and Bass, 2021). A recent report demonstrated significant daytime variation in multiple immune parameters in >300,000 participants in the UK Biobank, highlighting the diurnal nature of innate and adaptive immune responses (Wyse et al., 2021). Human lung diseases frequently show time-of-day variation in symptom severity and respiratory function and the circadian transcriptional activator BMAL1 has been shown to regulate respiratory inflammation (Ehlers et al., 2018; Ince et al., 2019). Influenza A virus infection of circadian-arrhythmic mice is associated with elevated inflammatory responses and a higher viral burden (Edgar et al., 2016; Sengupta et al., 2019). The time-of-day of influenza vaccination in elderly men affected antibody responses with higher titres noted in the morning (Phillips et al., 2008; Long et al., 2016). An additional influenza vaccination study reported that the time of sample collection rather than vaccination had a more significant effect on antibody responses (Kurupati et al., 2017). We and others have proposed a role for circadian signalling in regulating SARS-CoV-2 host immune responses and COVID-19 severity (Ray and Reddy, 2020; Maidstone et al., 2021; Sengupta et al., 2021). Clearly, it is important to assess whether the time of SARS-CoV-2 vaccination impacts host antibody responses.

In the UK, healthcare workers were identified as a priority group to receive SARS-CoV-2 vaccine starting in December 2020. At this time, the Alpha B.1.1.7 variant was the dominant circulating strain. As part of this initiative, data were collected on all asymptomatic staff members (Eyre et al., 2021; Lumley et al., 2021) in keeping with enhanced hospital infection prevention and control guidelines issued by the UK Department of Health and Social Care. Anonymised data were obtained from the Infections in Oxfordshire Research Database which has approvals from the National Research Ethics Service Committee South Central – Oxford C Research Ethics Committee (19/SC/0403), the Health Research Authority and the national Confidentiality Advisory Group (19/CAG/0144). Peripheral blood samples were collected during Dec 2020-Feb 2021 and were tested for anti-Spike (Abbott IgG assay) (Ainsworth et al., 2020) and anti-nucleocapsid (Abbott SARS-CoV-2 IgG anti-nucleocapsid assay) antibody levels. We analysed anti-Spike responses during the 2-10 weeks after vaccination. In this data set, 2190 people contributed one blood sample, 549 contributed two samples and 45 three or more samples (total of 3425 samples). Participants with evidence of prior SARS-CoV-2 infection (PCR for viral RNA or anti-nucleocapsid antibody), samples with anti-Spike responses <50 AU, and samples obtained after second vaccination were excluded.

Data from 2784 participants (Table 1A) were analyzed using linear mixed modelling to investigate the effects of time of vaccination on anti-Spike antibody levels. Variation between participants was modelled with fixed factors of time-of-day of vaccination (Time 1, 07:00-10:59; Time 2, 11:00-14:59; Time 3, 15:00-21:59) (Supplemental Figure 1), vaccine type (Pfizer, mRNA bnt162b2 or AstraZeneca, Adenoviral AZD1222), age group (16-29, 30-39, 40-49 or 50-74 years), sex, and the number of days post-vaccination. A B-spline transformation of days post-vaccination was used to model the non-linear pattern of anti-Spike responses (log10 transformed) (Supplemental Figure 2). This analysis allowed us to estimate the average anti-Spike levels in each participant group at 2 and 6 weeks post-vaccination (Figure 1).

**Table 1A.** Participant numbers.

|  | **Pfizer mRNA**<br>(Time 1/Time 2/Time 3) |  | **AstraZeneca Adenoviral**<br>(Time 1/Time 2/ Time 3) |  |
| --- | --- | --- | --- | --- |
| **Age (years)** | **Female** | **Male** | **Female** | **Male** |
| 16-29 | 90/143/163 | 18/26/26 | 39/54/53 | 11/12/10 |
| 30-39 | 100/146/149 | 30/46/40 | 38/44/34 | 10/7/8 |
| 40-49 | 120/160/170 | 17/36/42 | 43/56/43 | 8/11/8 |
| 50-74 | 127/152/199 | 24/26/38 | 68/52/59 | 7/4/7 |

**Figure 1:**
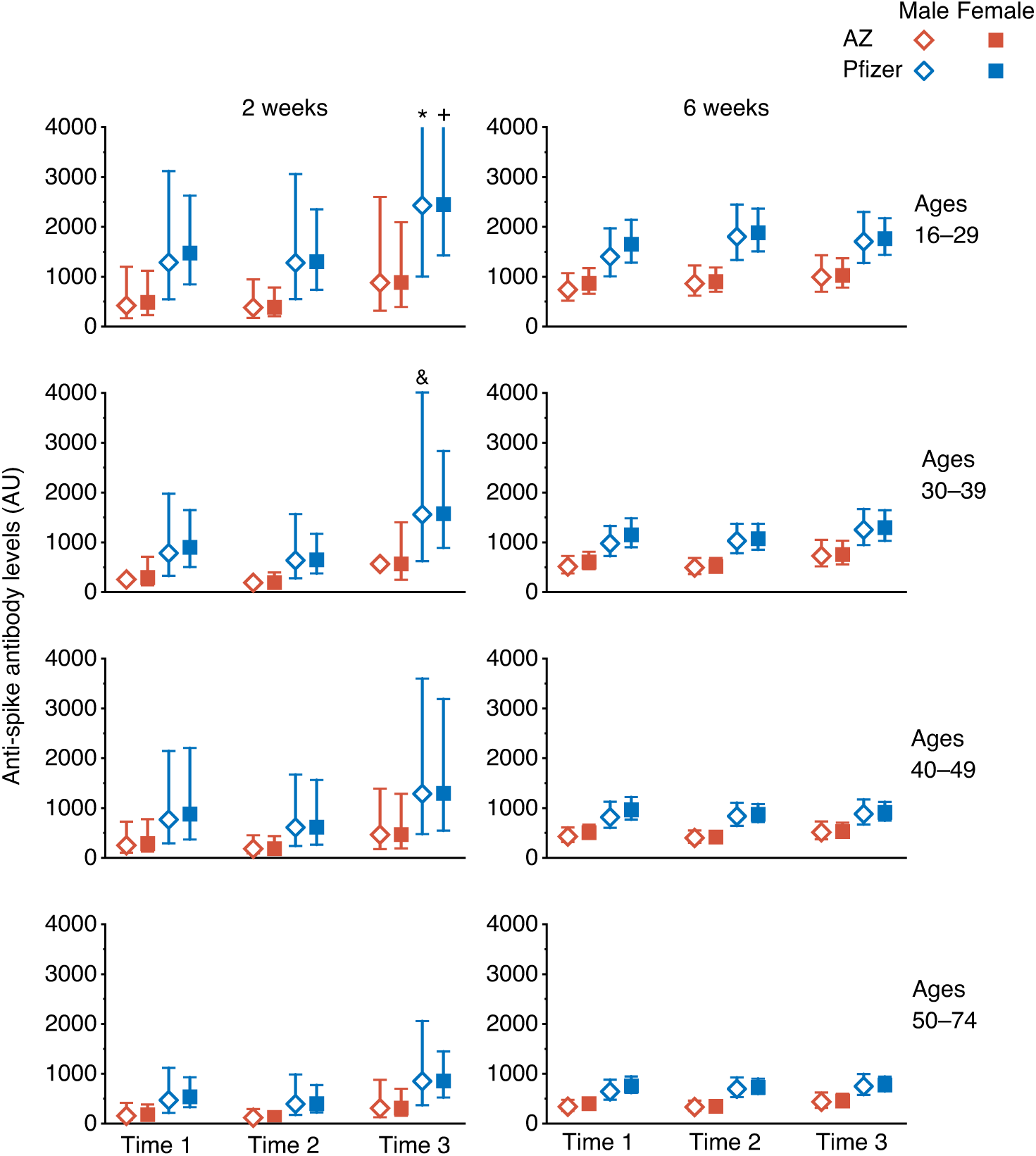
Estimated Anti-Spike antibody levels at 2 and 6 weeks after first SARS-CoV-2 vaccination, partitioned by age, sex, and time-of-day of vaccination (Time 1, 07:00-10:59; Time 2, 11:00-14:59; Time 3, 15:00-21:59). Mean value (symbol) with 95% confidence values (vertical line). † Three confidence intervals extend beyond the Y-axis limits (*4275; ^+^5995; ^&^4028).

Using a linear mixed-model approach, we found that anti-Spike responses were higher in those who were vaccinated later in the day (p=0.013), in those who received the Pfizer mRNA vaccine (p<0.0001), in women (p=0.013) and in younger participants (p<0.0001) (Table 1B). We observed significant interactions between days post-vaccination and vaccine type (p<0.0001) and age (p=0.032), but not with vaccine time (p=0.238). Analysing the data using two time intervals (before or after 1 pm) gave similar results. We did not observe a significant effect of time of day of sample collection (using the same time intervals as for vaccination times) (p=0.097), and this parameter was not included in the final model; results from the model including sample times are shown in Supplemental Table 1. Sixty seven samples gave values beneath the cut-off (<50) in the anti-Spike assay and were classified as “non-responders”, we found no significant association with the time-of-day of vaccination for these samples (linear mixed-effects logistic regression, p=0.23).

**Table 1B.** Type III tests of fixed effects from mixed effects model.

| **Effect** | **Num DF** | **F** | **Probability** |
| --- | --- | --- | --- |
| *Main Effects* |  |  |  |
| Vaccination Time<br>(Time 2, Time 3 vs. Time 1) ‡ | 2 | 4.33 | 0.0133 |
| Vaccine type<br>(AstraZeneca vs. Pfizer) | 1 | 148.31 | <0.0001 |
| Age<br>(30-39, 40-49, 50-74 vs.16-29) | 3 | 51.15 | <0.0001 |
| Sex<br>(Female vs. Male) | 1 | 6.16 | 0.0131 |
| Days post-vaccination | 6 | 18.78 | <0.0001 |
| *Interaction terms* |  |  |  |
| Days*Vaccination_Time | 12 | 1.26 | 0.2380 |
| Days*Vaccine type | 6 | 7.24 | <0.0001 |
| Days*Age | 18 | 1.70 | 0.0319 |
| Days*sex | 6 | 1.03 | 0.4010 |
| Vaccination_Time*Vaccine type | 2 | 1.22 | 0.2945 |
| Vaccination_Time*Age | 6 | 0.71 | 0.6446 |
| Vaccination_Time*Sex | 2 | 0.44 | 0.6412 |

DF= Degrees of Freedom.† For all F tests the denominator DF was 3359. ‡ For each F test, the fixed effect referent is the last term shown, the F and P values are the Type III tests of overall fixed effects.

Our analysis of 2784 healthcare workers reveals a significant effect of the time of vaccination on anti-Spike antibody levels following the administration of two alternative SARS-CoV-2 vaccines (mRNA or Adenovirus based). A recent report studying a small cohort of healthcare workers immunised with an inactivated SARS-CoV-2 vaccine in the morning (09:00 -11:00,n=33) or afternoon (15:00 - 17:00, n=30) showed increased B-cell responses and anti-Spike antibodies in participants vaccinated in the morning (Zhang et al., 2021). This contrasts with our observations and may reflect the use of an inactivated whole virus immunogen that will likely induce polytypic responses to a range of SARS-CoV-2 encoded proteins. Our observation contrasts with earlier studies in elderly men that reported higher anti-influenza titers in the morning (Phillips et al., 2008; Long et al., 2016). This may reflect differences between the cohorts studied, particularly with regard to immune status; we studied seronegative participants whereas responses to influenza vaccination will involve the stimulation of memory responses. Sample collection time in this study showed no significant association with anti-Spike levels, in contrast to previous reports (Kurupati et al., 2017; McNaughton et al., 2021). These data highlight the importance of recording the time of vaccination in clinical and research studies, and highlight the importance of considering time-of-day factors in future study designs that may reduce inter-individual variance and the number of participants needed to obtain statistical significance.

Additional studies are warranted to evaluate the circadian regulation of natural and vaccine-induced SARS-CoV-2 immunity. McNaughton and colleagues reported a diurnal variation in SARS-CoV-2 PCR test results, showing a 2-fold variation in Ct values implying higher viral RNA levels in the afternoon (McNaughton et al., 2021). These data are consistent with our recent study showing a role for the circadian component BMAL1 in regulating SARS-CoV-2 replication (Zhuang et al., 2021) that could influence the induction of host innate and adaptive responses.

It is worth noting that, despite the significant differences in anti-Spike levels detected in participants receiving Pfizer mRNA or AstraZeneca Adenoviral vaccines, both show comparable efficacies highlighting the robust nature of the host antibody response. Limitations of this retrospective observational study include: (i) relatively few participants had more than one anti-Spike antibody measurement, limiting our ability to study both longitudinal immune responses and the effect of time-of-day of sample collection; (ii) the health profiles of our healthcare workers may differ from the general population and no information was available on their medical or medication history, except that they had no prior infection with SARS-CoV-2 and were seronegative; (iii) there was limited serological sampling following second vaccination, precluding the analysis of time-of-day effects following a two-dose schedule; (iv) the extent to which anti-Spike levels are a correlate of clinical efficacy is not known; (v) the sleep and shift-work patterns of the participants, that are known to influence vaccine responses (Spiegel et al., 2002; Lange et al., 2003; Prather et al., 2021), were not available; and (vi) our cohort does not include children or high-risk groups, such as the elderly or immunocompromised. We recommend future studies address these limitations when documenting natural and vaccine-induced SARS-CoV-2 immune responses.

## Data Availability

All data produced in the present study are available upon reasonable request to the authors.

## Acknowledgements

We thank Helene Borrmann, Xiaodong Zhuang and Tanya Wilson for early discussions on this project; Tess Lambe and Merryn Voysey for their constructive comments on the paper. We also thank all OUH staff who participated in the staff testing program and the staff and medical students who ran the program. This work uses data provided by healthcare workers and collected by the UK’s National Health Service as part of their care and support. We thank all the people of Oxfordshire who contributed to the Infections in Oxfordshire Research Database. Research Database Team: L Butcher, H Boseley, C Crichton, O Freeman, J Gearing (community), R Harrington, M Landray, A Pal, TEA Peto, TP Quan, J Robinson (community), J Sellors, B Shine, AS Walker, D Waller. Patient and Public Panel: G Blower, C Mancey, P McLoughlin and B Nichols.

## Support

EBK is funded by NIH K24-HL105664, P01-AG009975 and R01-HL128538. JAM is funded by a Wellcome Investigator Award 200838/Z/16/Z, UK Medical Research Council project grant MR/R022011/1 and Chinese Academy of Medical Sciences Innovation Fund for Medical Science, China (grant number: 2018-I2M-2-002). WW is funded by NCATS Harvard Clinical and Translational Science Center grant 5UL1TR002541-02. DWE is a Robertson Foundation Fellow. DCA is funded by the NIHR Oxford Biomedical Research Centre. The report presents independent research. The views expressed in this publication are those of the authors and not necessarily those of the NHS, NIHR, or the UK Department of Health. PCM is funded by a Wellcome intermediate fellowship grant Ref 110110/Z/15/Z.

## Disclosures

WW has a consultancy for the National Sleep Foundation. PB has no relevant disclosures. DWE declares lecture fees from Gilead, outside the submitted work. EBK support from Gordon Research Conference, Sleep Research Society, Santa Fe institute, DGSM (German Sleep Society); consultancy for Circadian Therapeutics, National Sleep Foundation, Puerto Rico Science Technology Trust, Sanofi-Genzyme; partner owns Chronsulting. JAM has no relevant disclosures.

## Notes

### Author Declarations

All asymptomatic staff data collection and testing were part of enhanced hospital infection prevention and control measures instituted by the UK Department of Health and Social Care. Deidentified data were obtained from the Infections in Oxfordshire Research Database which has approvals from the National Research Ethics Service Committee South Central - Oxford C Research Ethics Committee (19/SC/0403), the Health Research Authority and the national Confidentiality Advisory Group (19/CAG/0144).

