## Supplemental materials for "Time of day of vaccination affects SARS-CoV-2 antibody responses in an observational study of healthcare workers"

**Supplemental Figure 1:**

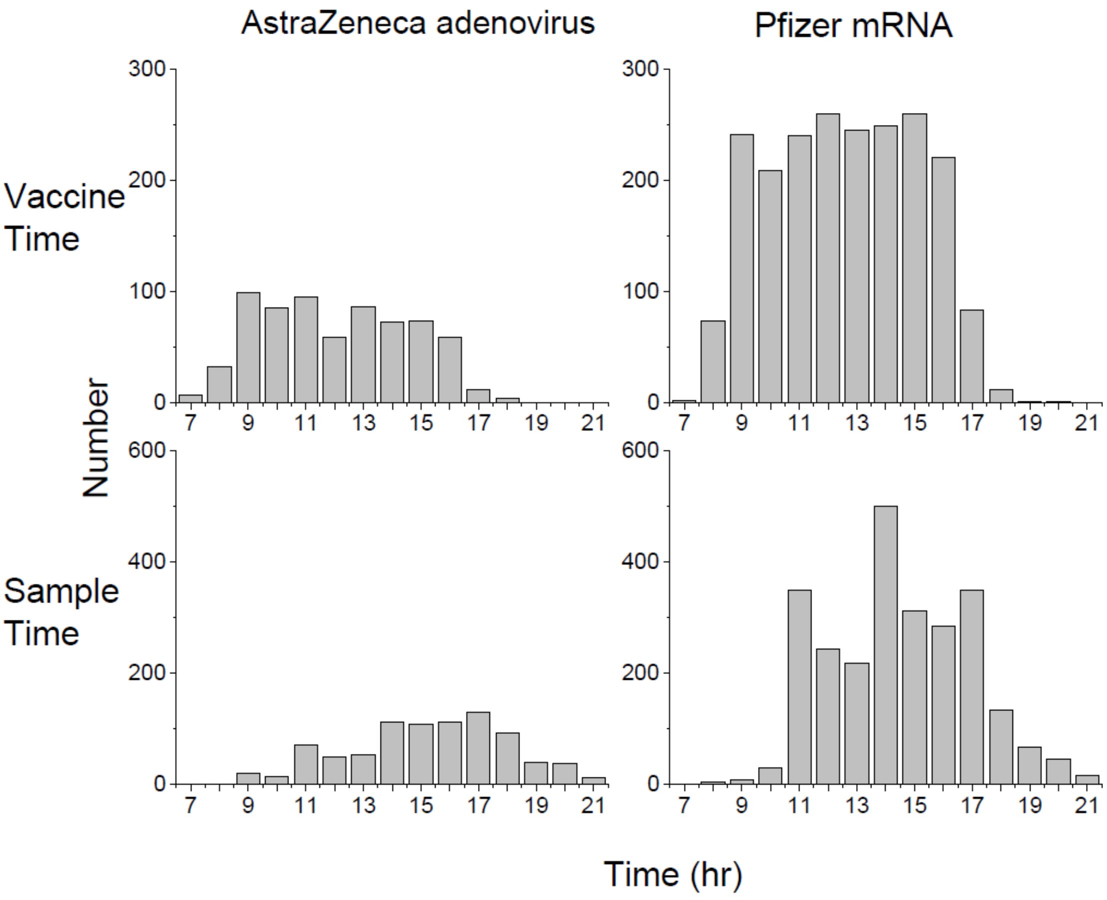


Histograms showing time (hour) of day of vaccinations (N=2784) and of sample collection (N=3425)

### Supplemental Figure 2

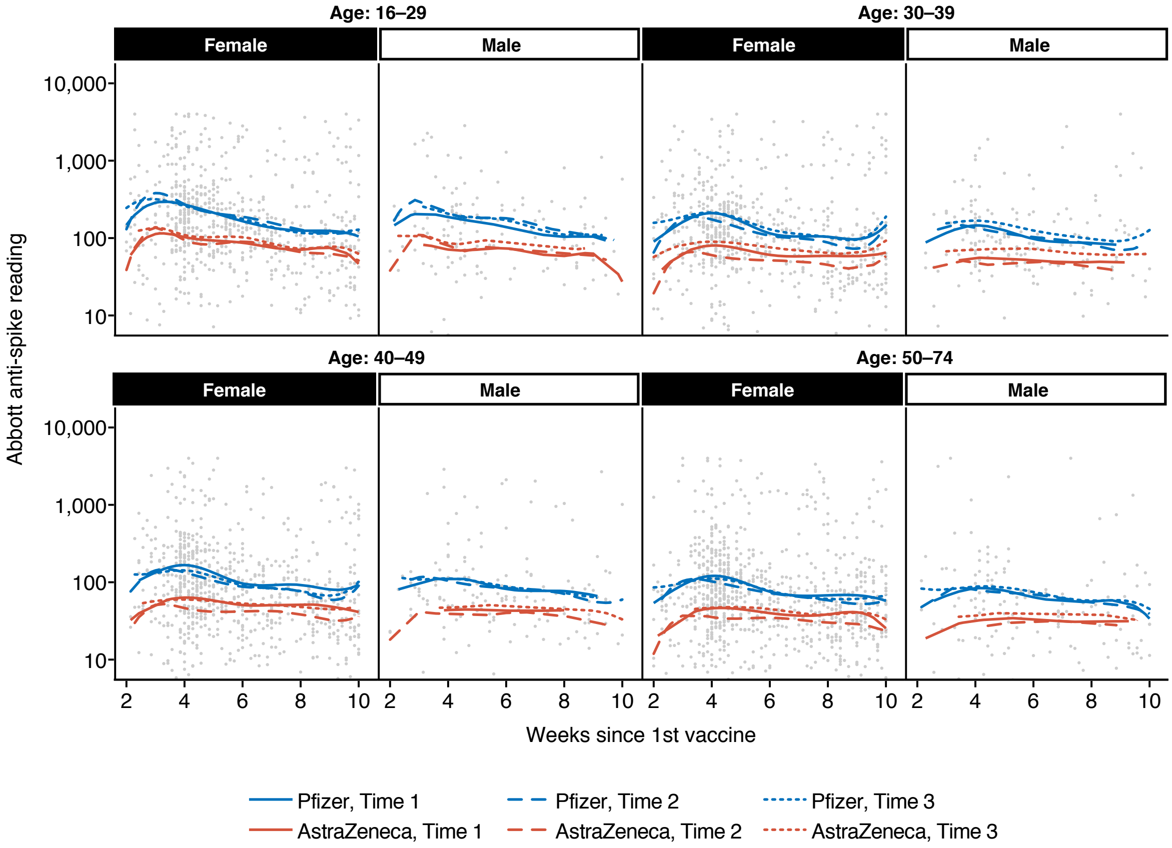


B-spline fit curves, including time-of-day of vaccination (Time 1, 07:00-10:59; Time 2, 11:00-14:59; Time 3, 15:00-21:59), vaccine type, age group, sex, and days post vaccination to $log_{10}$ (Anti-Spike antibody response) Data estimates are split by time-of-day of vaccination vaccine type (Pfizer mRNA, blue; AstraZeneca Adenovirus, red), age group, and sex (Female or Male).

**Supplemental Table 1: Type III tests of fixed effects from mixed effects model with sample time**

| Effect | Num DF | F | Probability |
|---|---|---|---|
| *Main Effects* | | | |
| Vaccination_Time (Time 1, Time 2, Time 3) ‡ | 2 | 4.40 | 0.0123 |
| Sample_Time (Time 1, Time 2, Time 3) ‡ | 2 | 2.34 | 0.0969 |
| Vaccine type (AstraZeneca vs. Pfizer) | 1 | 150.68 | <0.0001 |
| Age (30-39, 40-49, 50-74 vs.16-29) | 3 | 51.76 | <0.0001 |
| Sex (Female vs. Male) | 1 | 6.02 | 0.0142 |
| Days post-vaccination | 6 | 19.24 | <0.0001 |
| *Interaction terms* | | | |
| Days*Vaccination_Time | 6 | 1.28 | 0.2230 |
| Days*Vaccine type | 6 | 7.26 | <0.0001 |
| Days*Age | 18 | 1.73 | 0.0283 |
| Days*sex | 6 | 1.09 | 0.3684 |
| Vaccination_Time*Vaccine type | 1 | 1.23 | 0.2926 |
| Vaccination_Time*Age | 3 | 0.71 | 0.6387 |
| Vaccination_Time*Sex | 1 | 0.42 | 0.6604 |

Details of the linear mixed modeling are: Time of vaccination (Time 1, 07:00-10:59; Time 2, 11:00-14:59; Time 3, 15:00-20:59), vaccine type (Pfizer or AstraZeneca), age groups (from Table 1A), sex, and days post-vaccination were treated as fixed factors. A B-spline transformation of days post-vaccination was used to model the non-linear pattern of anti-Spike antibody responses (log10 transformed) post vaccination.
DF= Degrees of Freedom.† For all F tests the denominator DF was 3357. ‡ For each F test, the fixed effect referent is the last term shown, the F and P values are the Type III tests of overall fixed effects.
